# Impact of vaccination coverage and timing on recent yellow fever outbreaks in Africa: a modeling study

**DOI:** 10.64898/2026.11.11.26367845

**Authors:** Seth D. Judson, Jeffrey Pennington, Carlos Haring, Nicole Stephan, Mirava Yuson, Sayana Lee, Ernest Kenu, David Dowdy

## Abstract

**Background:** Multiple recent yellow fever (YF) outbreaks have occurred despite a highly effective vaccine. Increasing vaccination coverage and decreasing response times have been key components of the Eliminate Yellow Fever Epidemics (EYE) strategy. However, the combined impacts of these factors on YF outbreaks, especially since implementation of the EYE strategy in 2017, remain uncertain.

**Methods:** We developed mathematical models to simulate the effects of vaccination coverage and timing, calibrating them to recent YF outbreaks in Ghana (2021-2022) and South Sudan (2023-2024). We then projected YF morbidity and mortality under different EYE strategy thresholds for vaccination coverage and timing.

**Results:** In Ghana, where reactive vaccination was observed at 22 days with an estimated 70% baseline coverage, modeled delays of 60 and 120 days yielded 62 (95% uncertainty interval [UI] 30-119) and 281 (95% UI 59-927) additional severe cases, respectively. Increasing baseline vaccination coverage to 80% averted an estimated 20 severe cases (95% UI 14-25). In South Sudan, where reactive vaccination occurred at 61 days with estimated 50% baseline coverage, shortening the response time to 20 days and increasing baseline coverage to 80% averted an estimated 80 (95% UI 68-91) and 95 severe cases (95% UI 80-112) independently – or 103 severe cases (95% UI 85-120) if combined.

**Conclusion:** Increasing baseline YF vaccination coverage to at least 80% can avert substantial morbidity from YF outbreaks. Decreasing vaccination response time for reactive vaccination can provide additional benefit.

**Summary:** Gaps in immunization and delays in reactive vaccination contribute to yellow fever outbreaks, but specific impacts are unknown. This modeling analysis estimates the individual and combined effects of vaccination coverage and response timing on recent yellow fever outbreaks in Africa.

## Introduction

Yellow fever (YF) is a vaccine-preventable disease caused by YF virus, which is transmitted by mosquitoes in Africa and South America. In 2016 a yellow fever (YF) epidemic in Africa depleted the global supply of YF vaccines [1]. This led to the creation of the Eliminate Yellow Fever Epidemics (EYE) strategy by the World Health Organization (WHO) and Gavi [2]. The goal of the EYE strategy has been to eliminate YF epidemics by 2026 through increased investment and support in YF vaccination and laboratory diagnostic capacity [2]. It is estimated that over 336 million individuals have been vaccinated in Africa since the EYE strategy [3].

Despite this effort, there have continued to be recent YF outbreaks in multiple African and South American countries [3,4]. In 2024, 15 countries in Africa reported YF cases to the WHO, requiring vaccination interventions [3]. As the EYE strategy transitions to greater country-level ownership after 2026 [5], it is critical to assess its impact, as well as how both the coverage and timing of YF vaccination affect the trajectory of outbreaks.

A focus of the EYE strategy has been on increasing YF vaccination coverage and availability. There are three main YF vaccination strategies: routine childhood vaccination, preventative mass vaccination campaigns (PMVCs), and reactive vaccination campaigns. In YF-endemic countries, routine childhood vaccination for YF occurs usually at 9 months of age. Preventative mass vaccination campaigns are designed to increase YF vaccination coverage in endemic areas. Timely reactive vaccination during outbreaks has been a major component of the EYE strategy, emphasizing immediate YF vaccination using vaccines already available in-country followed by larger mass vaccination campaigns [2,6,7].

The International Coordinating Group of Vaccine Provision (ICG) manages the global YF vaccine stockpile [2]. Because the YF vaccine takes roughly a year to produce, the ICG stockpile is designed to ensure that sufficient doses are available at all times [2]. When a YF outbreak occurs, countries are recommended to promptly apply for YF vaccines. The WHO advises that there should be a maximum 10-day period between submission of the vaccine request and arrival [6]. Countries should then implement the reactive YF vaccination campaign within 10 days after receiving the vaccines [6]. Thus, a 20-day window is the approximate guideline for the time between YF outbreak detection and initiation of reactive mass vaccination.

The EYE strategy recommends a YF immunization coverage of “greater than 80%, with a 60-80% security threshold,” to prevent epidemics [2]. Additionally, the EYE strategy aimed to achieve at least 50% vaccination coverage in high-risk African countries via PMVCs [2]. There is a need to understand how these thresholds influence YF outbreaks in local contexts, as well as how vaccination timing and coverage synergistically influence YF outbreaks.

Given these questions and the upcoming transition of the EYE strategy, our study had three main goals. First, we developed mathematical models to assess the effects of immunization coverage and reactive vaccination timing on YF outbreaks. Second, we fit these models to recent outbreaks in Ghana (2021-2022) and South Sudan (2023-2024). Lastly, we simulated the effects of different YF immunization coverage and timing scenarios on projected outbreak size and mortality.

## Methods

### Modeling Framework

To model the effects of varying vaccination timing and coverage on YF outbreaks in African countries, we developed deterministic transmission models that incorporate stochastic surveillance reporting. The human (h) population was simulated using a Susceptible-Exposed-Infectious-Recovered (SEIR) framework, coupled with a Susceptible-Exposed-Infectious (SEI) model for the mosquito (v) population. This model framework was chosen because YF is an acute immunizing infection transmitted via mosquitoes, and it is assumed that infection and/or vaccination in humans provides lifelong immunity. For the purposes of analyzing outbreak scenarios and outcomes, we assumed that these were closed outbreaks (excluding human births, non-YF related deaths, and migration during the outbreaks). The underlying compartmental model framework is shown in Figure 1. The human compartments include susceptible (Sh), exposed (Eh), symptomatic/severe infected (Ih), asymptomatic/non-severe infected (Ah), toxic (Th), and recovered (Rh). The mosquito compartments include susceptible (Sv), exposed (Ev) and infected (Iv). The full model, parameter notation, and equations are included in the Appendix (Figure S1).

**Figure 1.**
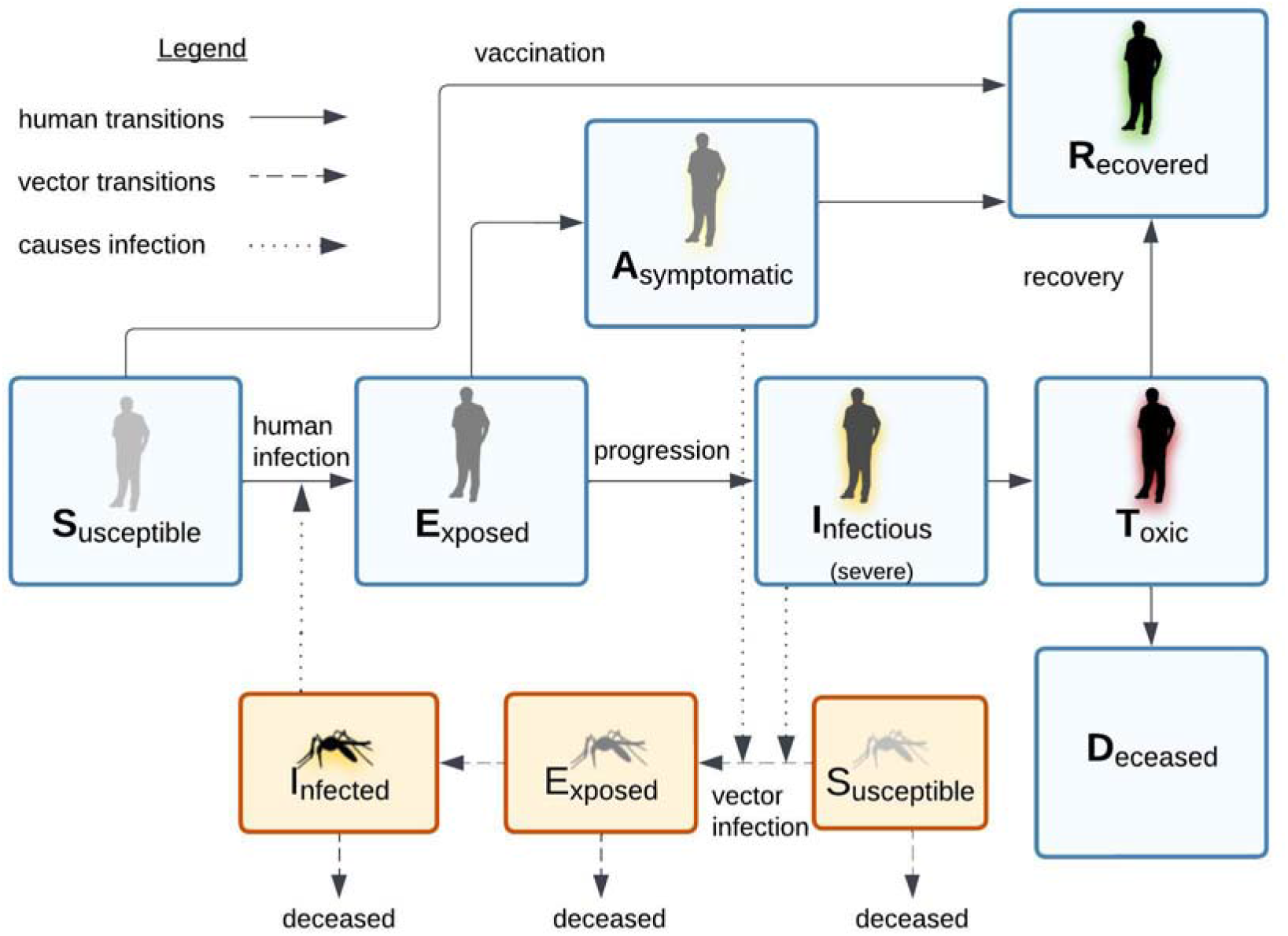
Yellow fever mechanistic model.

The compartments for the SEIR-SEI yellow fever (YF) model framework are shown in blue for humans and orange for mosquitoes. Susceptible mosquitoes are exposed by biting an infected human with asymptomatic or severe infection. These exposed mosquitoes go on to develop active infection and then can transmit to susceptible humans. There are three pathways for susceptible humans upon exposure to YF virus from an infected mosquito: 1. Those who are vaccinated prior to infection join the recovered compartment. 2. Those who are exposed and are asymptomatic/non-severe recover. 3. Those who are exposed and become severely infected and go on to the toxic state to either recover or die.

The model framework reflects separate pathways for humans who are exposed (Eh) to YF virus based on the force of infection (a function of the biting rate (*a*), transmission probability from vector to host (*b*), and ratio of infectious mosquitoes per human). Exposed individuals become infectious at an incubation rate (σh), and a proportion (δ) become severe infected individuals (Ih), while the remaining are asymptomatic/non-severe cases (Ah). The severe cases transition to the toxic state (Th) and then either die (Dh) or recover (Rh). We included a transition rate (η) between infected to toxic, which differs from the rate to recovery [8]. The asymptomatic/non-severe cases recover at rate γh. These separate pathways reflect the fact that YF surveillance systems in African countries rely on passive reporting and detect primarily severe/symptomatic cases [9]; a similar framework was used to model the 2016 Angola YF outbreak [10]. Model calibration and projections were fitted to the number of severe/symptomatic cases (Ih), as this compartment most closely reflects the observed surveillance data. Hereafter, the term “cases” refers to these severe/symptomatic individuals.

We used a partially observed Markov process (POMP) modeling framework [11], given that infected cases are only partially captured by the surveillance data. Because our goal was to create a model framework that could be flexible enough to use in different outbreak settings, we intentionally adopted a simplified, transparent modeling framework requiring estimation of a minimum number of parameters. We made an *a priori* assumption that outbreaks began with a single infected human (Ih = 1). Given that YF cases are likely to be under-ascertained in many African countries, we estimated the reporting ratio (ρ), defined as (ascertained cases)/(total cases). Additionally, we estimated the mosquito-to-human ratio (m) and the initial number of infected mosquitoes (Iv) as independent parameters in the model. The total mosquito population (Nv) was modeled as being at steady-state for the outbreak period, but a temperature-dependent mortality rate (μ_v_) balanced by an equivalent recruitment rate was used to simulate vector turnover (Appendix).

### Model Parameters

The values for the fixed and estimated parameters in the models are shown in Table 1. Since *Aedes aegypti* is the mosquito species responsible for YF epidemics in Africa [12], we used *Aedes aegypti*-specific parameters. We captured seasonal dynamics by using temperature-dependent functions for biting rate (*a*), mortality rate (μv), and vector incubation rate (σv), based on functions and mean parameter estimates for *Aedes aegypti* and YF (Appendix, Table S1) [13–15].

**Table 1.** Fixed Parameters.

| Parameter | Notation | Value/Range | Unit | Reference |
|---|---|---|---|---|
| Total human population | Nh | 653,266 (Ghana Savannah Region)<br>663,235 (South Sudan, Western Equatoria Region) | people | Ghana population: 2021 DHIMS2<br>South Sudan 2023: UNFPA |
| Initial number of infected humans | Ih | 1 | people |  |
| Human incubation rate | σh | 0.25 | days | [16] |
| Vector incubation rate | σv | 0.11-0.13 (Ghana)<br>0.09-0.13 (South Sudan) | days | [17] |
| Human recovery rate | γh | 0.25 | days | [10] |
| Proportion of infections that are symptomatic/severe | δ | 0.12 |  | [9] |
| Transition rate from infected to toxic | η | 0.5 | days | [8] |
| Case fatality ratio for severe YF | θ | 0.095 |  | [18] |
| Transition rate from toxic | k | 0.125 | days | [10] |
| Mosquito biting rate | a | 0.38-0.44 (Ghana)<br>0.35-0.43 (South Sudan) | per vector day | [15] |
| Transmission probability from vector to host | b | 0.4 | per bite | [10] |
| Transmission probability from host to vector | c | 0.5 | per bite | [10] |
| Relative infectivity of non-severe cases | ξ | 0.5 |  | [10] |
| Mosquito death rate | μv | 0.037-0.044 (Ghana)<br>0.035-0.042 (South Sudan) |  | [14] |
| Vaccination coverage |  | 0.5, 0.6, 0.8 |  | [2] |
| Vaccination rate during reactive vaccination campaign | Vt | 0.079 | Proportion vaccinated per day | [18] |
| Vaccination start delay |  | 20, 60, 120 | days | [18] |

We created two versions of the model for different YF outbreaks (2021-2022 Ghana and 2023-2024 South Sudan). These were deemed appropriate comparators given that both outbreaks occurred post-EYE strategy in regions with similar population sizes. The fixed parameters remained the same for both models except for the total human populations and temperature-dependent mosquito parameters. Calibration of the model for each outbreak setting is described further below and in the Appendix (Figures S2-S7).

For the Ghana version of the model, we used YF case data only from the Savannah region, which is where the outbreak originated and most cases occurred (46/71 cases)[19,20]. We did this to account for the fact that climatic and mosquito dynamics vary between the northern savannah and southern forested zones in Ghana. Additionally, through narrowing the spatial scale, the homogeneous mixing assumption of the model was more closely met. The 2021-2022 YF case data were obtained from the Ghana Health Service as previously described [20,21]. The average mean temperature in the Savannah region for each month of the outbreak was extracted from TerraClimate using Google Earth Engine. These mean temperature values were then included in temperature-dependent mosquito functions. Population data in the Savannah region were obtained from the 2021 DHIMS2 in Ghana. The YF vaccination coverage in the Savannah region at the start of the outbreak was estimated at 70%, combining routine and supplemental vaccination campaign data as previously described [20]. Routine childhood immunization for YF has been implemented in Ghana since 1992.

The 2023-2024 South Sudan outbreak occurred in the Western Equatoria region, and monthly mean temperature data during the outbreak period was similarly obtained and included as above. Population data for Western Equatoria were obtained from the 2023 UNFPA (https://data.humdata.org/dataset/cod-ps-ssd). In contrast to Ghana, routine childhood immunization for YF and regional coverage estimates were unavailable for South Sudan. Therefore, we used the WHO/UNICEF estimates from Sudan, which estimated 50% coverage in 2023 [22]. The South Sudan 2023-2024 YF case data were obtained from a public situation report [23].

Reactive vaccination campaigns occurred in response to both the Ghana and South Sudan outbreaks. We used data from a previously described literature review on YF outbreaks in Africa for the case fatality ratio (CFR) for severe YF (total deaths reported/total cases reported) and overall average vaccination rate (vt) [18]. The CFR including all deaths and all reported cases was chosen because the data from the Ghana outbreak included probable/confirmed cases, while the South Sudan outbreak included suspected cases. A delay of 10 days from YF vaccination to immunity was included in the models based on international guidelines [24].

### Model Calibration

We calibrated the reporting ratio (ρ), number of initial infected mosquitoes (Iv), and the mosquito-to-human ratio (m) for the Ghana and South Sudan outbreaks by fitting model-estimated cases to observed daily YF cases, assuming estimated vaccination coverage and reactive vaccination timing for each outbreak as described above. We calibrated the size of the Ih compartment to the number of observed cases on a daily basis, using data on symptom onset from both outbreaks. We implemented our model using the pomp package in R [11].

We used likelihood-based parameter estimation and sampled the parameter space using Latin Hypercube Sampling (LHS) with 10,000 iterations. The model was calibrated by minimizing the negative log-likelihood (NLL), assuming the observed cases for each outbreak day followed independent Poisson distributions (evaluated using the *dmeasure* function). To account for uncertainty, we first selected all parameter sets within 10 NLL units of the best fitting parameter set (Figure S4-S5). To perform the scenario analyses of different vaccination coverages and response times, we took likelihood-weighted samples of these parameter sets (4,400 per outbreak) and used the outputs from those likelihood-weighted samples to estimate the size of outbreaks under different scenarios (Appendix). Likelihood weights were based on the fit of deterministic model projections to each day’s case count, under the assumption that these case counts represented independent, Poisson-distributed variables, scaled by the reporting fraction (ρ).

The prior and posterior distributions of ρ, Iv, and m – including posterior mean values and 95% uncertainty intervals (UIs) for both Ghana and South Sudan – are shown in Table 2. For the mosquito-to-human ratio (m), we initially set the prior range from 1-4 [25]. We subsequently expanded the range of m to an upper limit of 8, the maximum value that we found reported for *Aedes aegypti* [26], based on initial calibration results in which the posterior range clustered near the upper bound of the prior range.

**Table 2.** Estimated Parameters.

| **Parameter** | **Notation** | **Prior Distribution** | **Posterior mean, 95% UI (Ghana)** | **Posterior mean, 95% UI (South Sudan)_** | **Reference for prior** |
|---|---|---|---|---|---|
| Mosquito-to-human ratio | m | 1-8, discrete uniform | 4.15 (1.82-6.97) | 4.47 (3.53-5.49) | [25,26] |
| Initial number of infected mosquitoes | Iv | 20-500, discrete uniform | 333 (173-490) | 203 (71-439) |  |
| Case reporting probability | ρ | 0.2-1, uniform | 0.66 (0.34-0.98) | 0.50 (0.21-0.94) |  |

### Scenario Analysis

Once the models were calibrated to the Ghana and South Sudan outbreaks, we conducted scenario analyses to compare the effects of vaccination coverage and timing on the primary outcome of expected severe YF cases. We also evaluated a secondary outcome of YF deaths. For vaccination coverage, we used WHO thresholds of 50%, 60%, and 80%, and for vaccination delays we used the WHO guideline (20 days) and the approximate 25th and 75th percentiles (60 days and 120 days) from our prior literature review [18]. We ran these different scenarios on models separately calibrated to the Ghana and South Sudan outbreaks. All statistical analyses and models were conducted using R (version 4.5.3).

### Ethical Considerations

This study was deemed exempt from institutional review board (IRB) review by the University of California, Los Angeles IRB (IRB-25-1440), and informed consent was waived.

## Results

### Model calibration

During the 2021-2022 Ghana Savannah outbreak, there were 46 cases (45 with date of symptom onset available) and 6 deaths reported. There was a delay of 22 days from the index case symptom onset to reactive local vaccination, and 51 days to reactive mass vaccination. In the 2023-2024 South Sudan outbreak, there were 110 cases and 6 deaths reported, with a delay of 61 days between the index case and mass reactive vaccination. The distributions of observed versus modeled cumulative cases, assuming 70% baseline vaccination coverage in Ghana and 50% in South Sudan, are shown in Figure 2.

**Figure 2.**
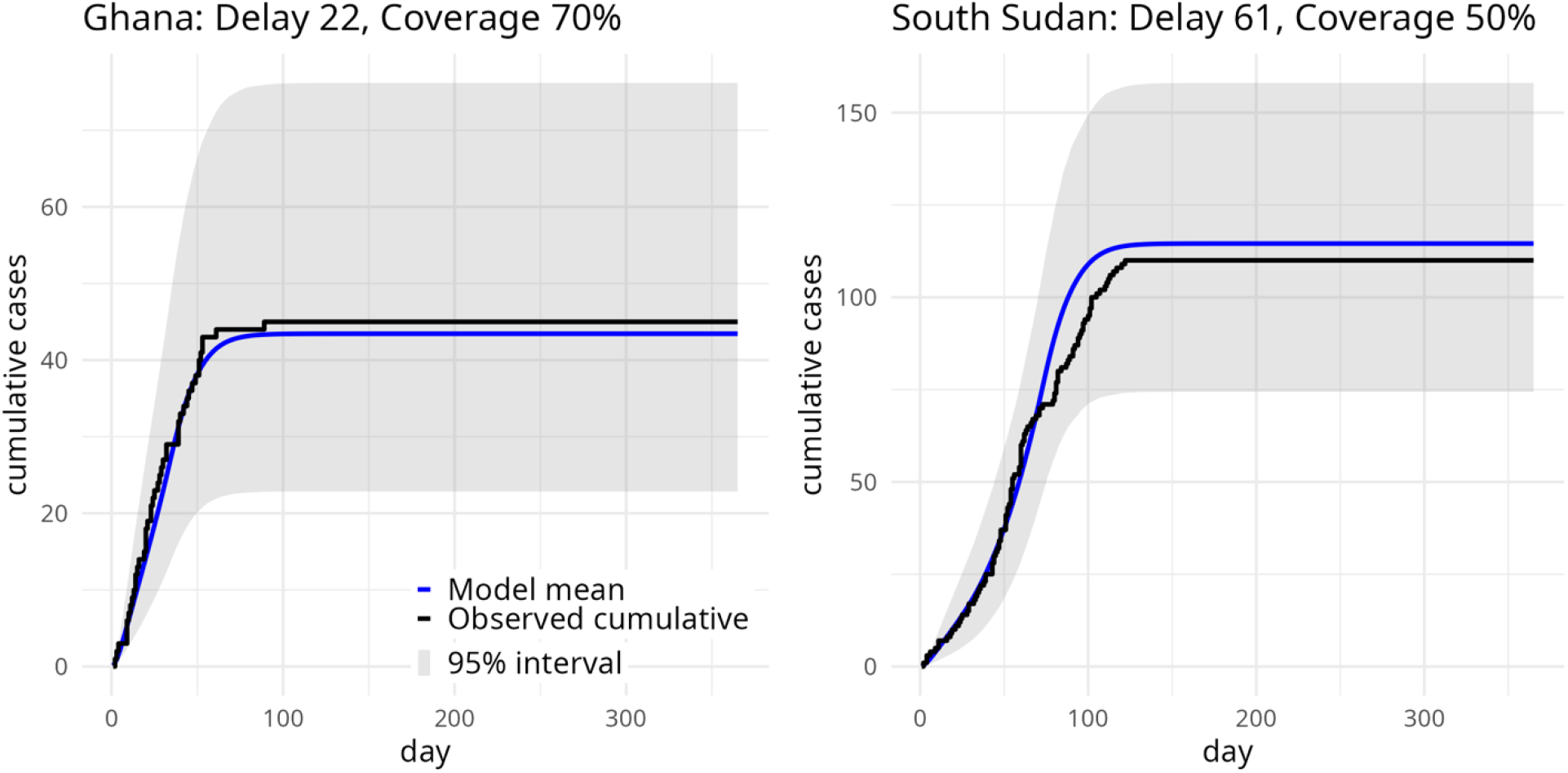
Observed vs modeled cumulative yellow fever cases in Ghana and South Sudan.

The figure shows the model calibrated to two yellow fever outbreaks: 2021-2022 Ghana (Panel A) and 2023-2024 South Sudan (Panel B). The observed number of cumulative daily cases are shown in black, and the model-estimated numbers of severe/toxic cases are shown in blue, with shaded regions denoting 95% uncertainty intervals).

### Effect of delayed vaccination

Using the model calibrated to the Ghana outbreak, extending the delay between outbreak onset and reactive vaccination from 22 to 60 or 120 days was estimated to result in an additional 62 (95% UI 30-119) and 281 cases (95% UI 59-927), respectively (Table S2). In comparison, using the model calibrated to South Sudan, if time to reactive vaccination were shortened from 61 to 20 days, an estimated 80 cases (95% UI 68-91) would have been averted (Tables S5).

Figure 3 shows the simulated trajectories based on the Ghana model using delays of 20, 60, and 120 days and 50%, 60%, and 80% vaccination coverage. Estimated cumulative cases and deaths for each scenario are given in the Appendix (Tables S2-S4 for Ghana; Tables S5-S7 and Figure S8 for South Sudan).

**Figure 3.**
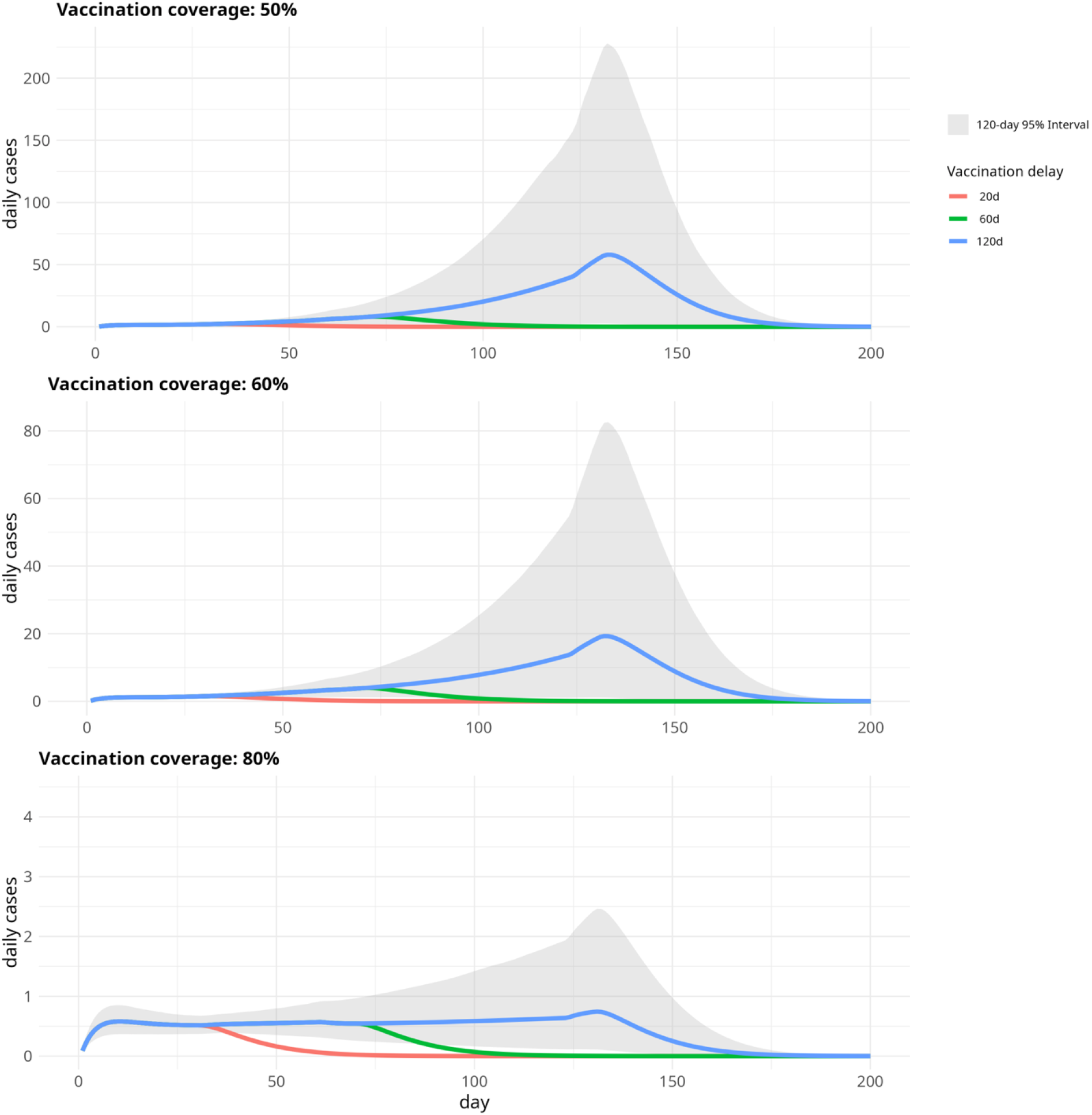
Estimated counterfactual yellow fever outbreak trajectories based on vaccination coverage and timing in Ghana, 2021-2022.

The figure shows estimated severe/symptomatic yellow fever cases in counterfactual scenarios assuming vaccination coverage of 50%, 60%, and 80%. The red line in each panel indicates the estimated epidemic assuming a delay of 20 days to reactive vaccination, versus 60 days (green line) and 120 days (blue line). The shaded grey region represents the 95% uncertainty interval under the 120-day delay scenario (which overlaps with the other scenarios of vaccination delay until the time of simulated reactive vaccination).

### Effect of vaccination coverage

In the Ghana model, increasing vaccination coverage from 70 to 80% was predicted to avert an estimated 20 cases (95% UI 14-25) (Table S3), while in the South Sudan setting, increased vaccination coverage from 50 to 80% was estimated to avert 95 cases (95% UI 80-112) (Table S6). The relative estimated impacts of vaccination timing and coverage are shown in Figure 4 (Ghana) and Figure S9 (South Sudan).

**Figure 4.**
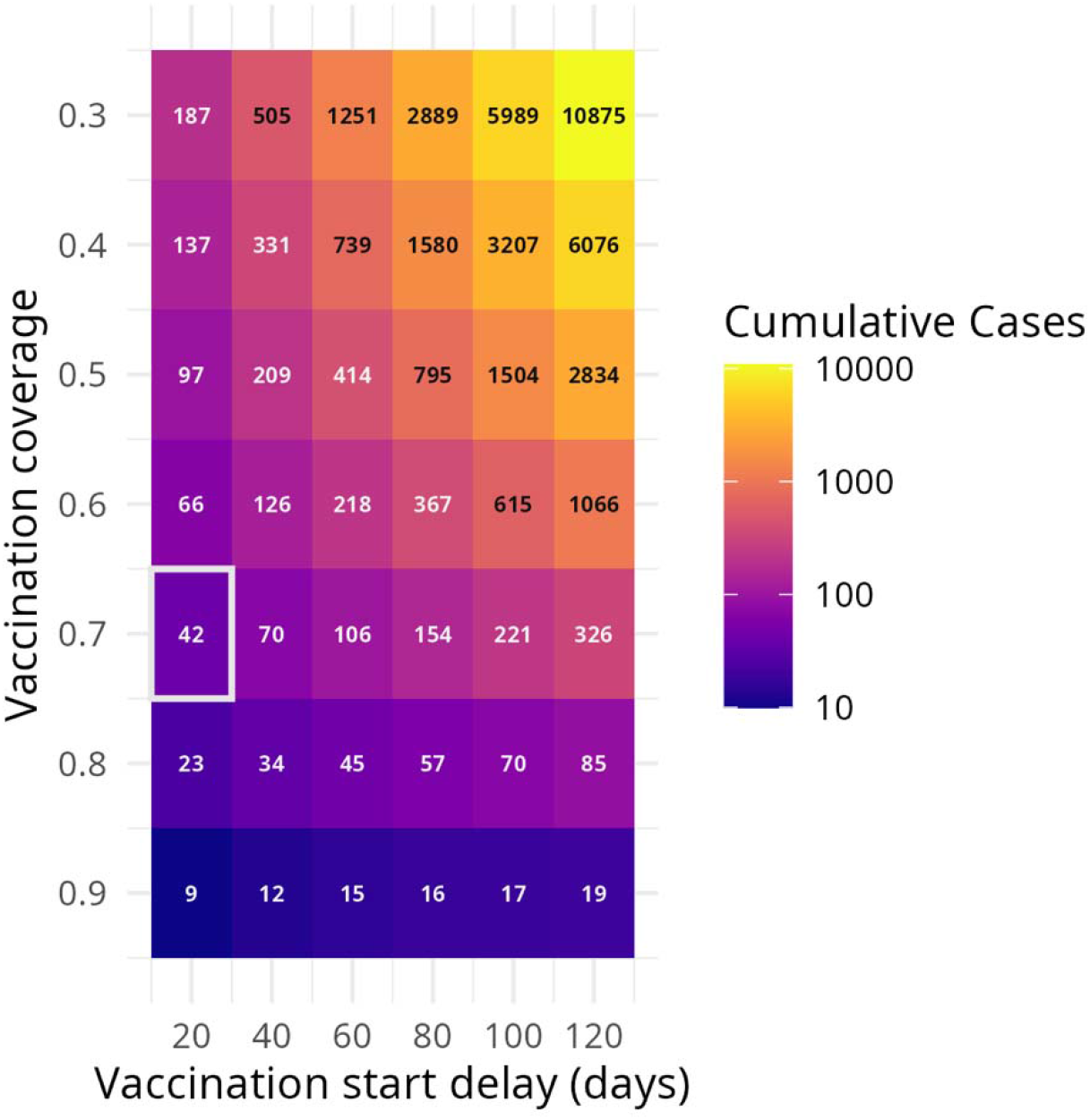
Estimated impact of vaccination coverage and timing of reactive vaccination in the 2021-2022 Ghana yellow fever outbreak.

The heatmap depicts the estimated number of cumulative symptomatic/severe yellow fever cases during the Ghana 2021-2022 outbreak, under counterfactual scenarios of baseline vaccination coverage and delay to start of reactive vaccination. Color intensity indicates the mean number of estimated cases on a logarithmic scale. The scenario closest to the observed epidemic (70% baseline vaccination coverage, 22-day delay, 46 observed cases) is outlined in white.

## Discussion

Since the EYE strategy was first implemented in 2017, there has been progress towards eliminating YF epidemics. However, recent YF events, including an epidemic in the Colombian Andes, highlight persisting gaps in immunization coverage in areas previously thought to be low risk [27]. Furthermore, there have been significant delays in reactive vaccination during recent outbreaks in Africa [18]. While previous studies have evaluated the effects of YF vaccination coverage and delays separately, here we provide a framework to quantify the impact of both factors on YF outbreaks.

We found that the recent outbreaks in Ghana (2021-2022) and South Sudan (2023-2024) are useful archetypes for modeling the effects of YF immunization. Ghana had a timely local YF vaccination campaign and regional vaccination coverage near the EYE threshold of 80%. Meanwhile, South Sudan had lower vaccination coverage, longer delays to reactive vaccination, and likely lower detection/reporting of cases – and an observed outbreak size approximately 2.5 times as large. In both scenarios, our models estimate that increasing baseline immunization coverage and decreasing the time to reactive vaccination would have averted a substantial number of cases and deaths. Furthermore, our models suggest that baseline vaccination coverage and reactive vaccination timing act non-linearly to influence outbreak size – at any given level of baseline coverage, reducing the time to reactive vaccination averts additional morbidity, and vice versa.

When considering the critical vaccination threshold, we found that the recommended threshold of 80% was effective at mitigating large-scale outbreaks. However, reaching the 80% threshold (or the 60-80% security range) did not prevent expansion in all circumstances, especially if reactive vaccination was delayed. This finding aligns with a recent study showing that 80% coverage was effective at reducing the probability of YF outbreaks, yet risk remained in regions with transmission variability and vaccination coverage uncertainty [28]. Likewise, our findings align with a modeling study of the 2016 YF epidemic in Angola which similarly found that delayed vaccination by 60 days would have increased cases and deaths roughly 3-fold [10]. As depicted by Figure 4, both WHO targets of 80% YF vaccination coverage and 20 days to reactive vaccination are effective measures for outbreak control – and achieving both targets simultaneously can greatly reduce the size of potential outbreaks when they occur.

These findings have important implications for future YF vaccination strategies. Both increasing immunization coverage and decreasing the time to reactive vaccination are essential to mitigating outbreaks. In an ideal scenario, both strategies should be emphasized. However, in the setting of resource constraints, it is important to consider how countries may prioritize resources. Our findings demonstrate that baseline YF immunity influences how much response timing matters. Therefore, a tailored approach to vaccination coverage and response timing may improve resource allocation. The modeling framework in this study provides a data-driven approach for future country-specific cost-effectiveness analyses, which could guide policymakers on whether to invest more in routine immunization vs response infrastructure (i.e. diagnostics and vaccine availability) depending on baseline immunization coverage.

There are limitations to this analysis. To make the models more generalizable and capture the acute phase of outbreaks, we modeled the human population as closed and the mosquito population as being at equilibrium. Likewise, we assumed homogeneous mixing and used estimated vaccination coverages, which may affect total trajectory estimates. For instance, because the Ghana outbreak scenario had high estimated vaccination coverage (70%) and a fast response, sustaining transmission required a greater force of infection, which may overestimate outbreak size in scenarios with lower coverage and longer delays. We also assumed lifelong immunity from prior YF vaccination. Despite these limitations, our approach offers a framework for evaluating the synergies and tradeoffs between YF vaccination coverage and response timing.

Overall, the EYE strategy has made significant progress. Both investment in increased vaccination coverage and timely response have averted YF cases and deaths. However, YF outbreaks continue to occur, including in areas previously deemed as low risk. Identifying gaps in vaccination coverage, as well as delays in diagnosis and response, will be key to targeted prevention strategies. Modeling frameworks such as this can help decision-makers evaluate the relative benefits of these strategies as we enter the next phase towards eliminating YF epidemics.

## Data availability

Data from this study and R scripts for the analysis are available at: https://github.com/Judson-Lab/YF_Scenario_Model_Africa and https://doi.org/10.6084/m9.figshare.34983865

## Acknowledgements

Research reported in this publication was supported by the National Institute of Allergy and Infectious Diseases of the National Institutes of Health under Award Number K08AI190125 to SDJ. The content is solely the responsibility of the authors and does not necessarily represent the official views of the National Institutes of Health. The funders had no role in study design, data collection and analysis, decision to publish, or preparation of the manuscript.

## Author contributions

Conceptualization: SDJ; Data curation: SDJ, NS, MY, CH, SL, EK; Formal Analysis: SDJ, JP, DD; Funding acquisition: SDJ; Methodology: SDJ, JP, DD; Writing – original draft: SDJ; Writing – review & editing: SDJ, JP, CH, NS, MY, SL, EK, DD

