## Appendix for "Impact of vaccination coverage and timing on recent yellow fever outbreaks in Africa: a modeling study"

**Table of Contents**

**1. Modeling framework**

*Figure S1. Yellow fever mechanistic model with parameters*

*Equations for yellow fever mechanistic model*

*Table S1. Temperature-dependent parameters and functions*

**2. Calibration**

*Figure S2. Ghana 2021-2022 outbreak epidemiological curve*

*Figure S3. South Sudan 2023-2024 outbreak epidemiological curve*

*Figure S4. Distribution of estimated parameters for Ghana outbreak model calibration*

*Figure S5. Distribution of estimated parameters for South Sudan outbreak model calibration*

*Figure S6. Observed vs modeled daily yellow fever cases in Ghana 2021-2022*

*Figure S7. Observed vs modeled daily yellow fever cases in South Sudan 2023-2024*

**3. Scenario Analysis**

*Table S2. Ghana outbreak cases and deaths, observed vs simulated timing delays*

*Table S3. Ghana outbreak cases and deaths, observed vs simulated vaccination coverage*

*Table S4. Ghana outbreak cases and deaths, observed vs simulated vaccination coverage and delays*

*Table S5. South Sudan outbreak cases and deaths, observed vs simulated timing delays*

*Table S6. South Sudan outbreak cases and deaths, observed vs simulated vaccination coverage*

*Table S7. South Sudan outbreak cases and deaths, observed vs simulated vaccination coverage and delays*

*Figure S8. Projected yellow fever outbreak trajectories based on vaccination coverage and timing, South Sudan*

*Figure S9. Heatmap of vaccination coverage vs timing on yellow fever outbreak size, South Sudan*

1. Modeling framework

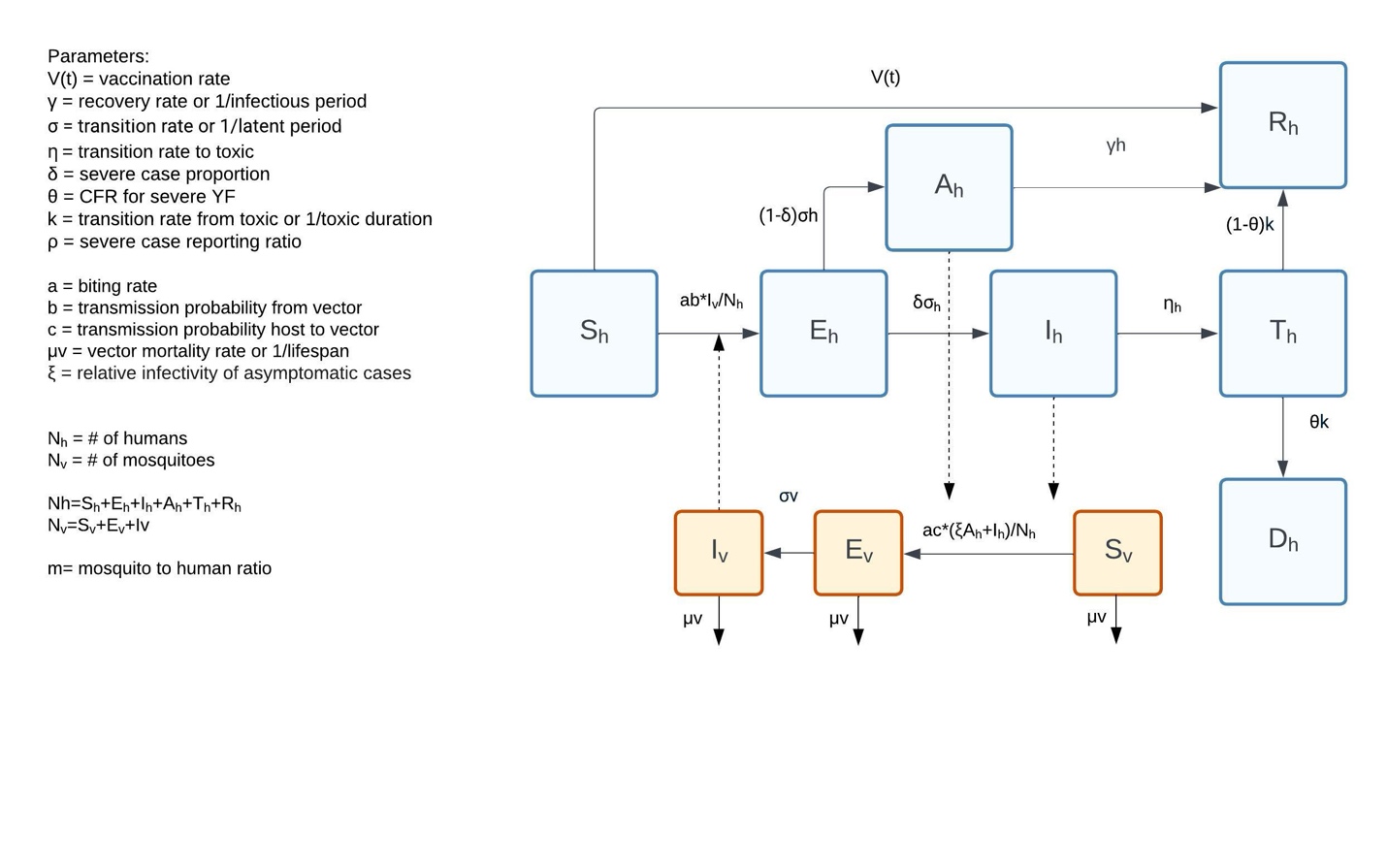
Figure S1. Yellow fever mechanistic model with parameters

The compartments and parameters for the yellow fever SEIR-SEI transmission model are shown. The human population compartments are depicted in blue and the mosquito compartments are orange. There are three pathways for susceptible humans: 1. Those who are vaccinated prior to infection and join the recovered compartment (Rh). 2. Those who are exposed (Eh) and are asymptomatic/non-severe (Ah) and recover. 3. Those who are exposed and become severely infected (Ih) and go on to the toxic state (Th) and either recover or die (Dh). The susceptible mosquitoes (Sv) become exposed (Ev) by biting an infected human with asymptomatic or severe infection. These exposed mosquitoes go on to develop active infection (Iv) and then can transmit to susceptible humans.

Equations for yellow fever mechanistic model

Human equations:

$$1.Nh=Sh+Eh+Ih+Ah+Th+Rh$$

$$2.\lambda h=\left( a*b*Iv \right)/{Nh}$$

$$3.\frac{dSh}{dt}=-\lambda h*Sh-\left( vt*Sh \right)$$

$$4.\frac{dEh}{dt}=\lambda h*Sh-\left( \sigma h*Eh \right)$$

$$5.\frac{dIh}{dt}=\delta*\sigma h*Eh-\left( \eta*Ih \right)$$

$$6.\frac{dAh}{dt}=\left( 1-\delta\right)*\sigma h*Eh-\left( \gamma h*Ah \right)$$

$$7.\frac{dTh}{dt}=\eta*Ih-\left( k*Th \right)$$

$$8.\frac{dRh}{dt}=\left( \gamma h*Ah \right)+\left( \left( 1-\theta\right)*k*Th \right)+\left( vt*Sh \right)$$

$$9.\frac{dDh}{dt}=\theta*k*Th$$

Mosquito equations:

$$1.Nv=Sv+Ev+Iv$$

$$2.Nv=m*Nh$$

$$3.\lambda v={a*c*\left( \left( \xi*Ah \right)+Ih \right)}/{Nh}$$

$$4.\frac{dSv}{dt}=\mu v*Nv-\lambda v*Sv-\mu v*Sv$$

$$5.\frac{dEv}{dt}=\lambda v*Sv-\left( \sigma v+\mu v \right)*Ev$$

$$6. \frac{dIv}{dt}=\sigma v*Ev-\mu v*Iv$$

*Temperature-dependent functions*

Temperature suitability functions were created based on prior estimates for *Aedes aegypti* biting rate (*a*), mortality (μ_v_), and yellow fever virus incubation rate (*σ*_v_). These functions were also combined to create an overall temperature suitability index for both Ghana and South Sudan settings. R scripts and data are available at:

https://github.com/Judson-Lab/YF_Scenario_Model_Africa

Table S1. Temperature-dependent parameters and functions

| Temperature -dependent parameter | Function | Scaling constant (c) | Thermal minimum (Tmin) | Thermal maximum (Tmax) | Reference |
| --- | --- | --- | --- | --- | --- |
| *a* (biting rate) | Brière | 0.000272 | 13.35 | 40.13 | [1] |
| μ_v_ (mortality rate) | Quadratic | 0.148 | 9.16 | 37.73 | [2] |
| *σ*_v_ (incubation rate) | Brière | 0.0001 | 17.67 | 42.1 | [3] |

1.	Gaythorpe KAM, Hamlet A, Jean K, et al. The global burden of yellow fever. eLife. eLife Sciences Publications Ltd; **2021**; 10.

2.	Mordecai EA, Cohen JM, Evans MV, et al. Detecting the impact of temperature on transmission of Zika, dengue, and chikungunya using mechanistic models. PLOS Neglected Tropical Diseases. Public Library of Science; **2017**; 11(4):e0005568.

3.	Gaythorpe KAM, Hamlet A, Cibrelus L, Garske T, Ferguson NM. The effect of climate change on yellow fever disease burden in Africa. eLife. eLife Sciences Publications Ltd; **2020**; 9:1–27.

2. Calibration

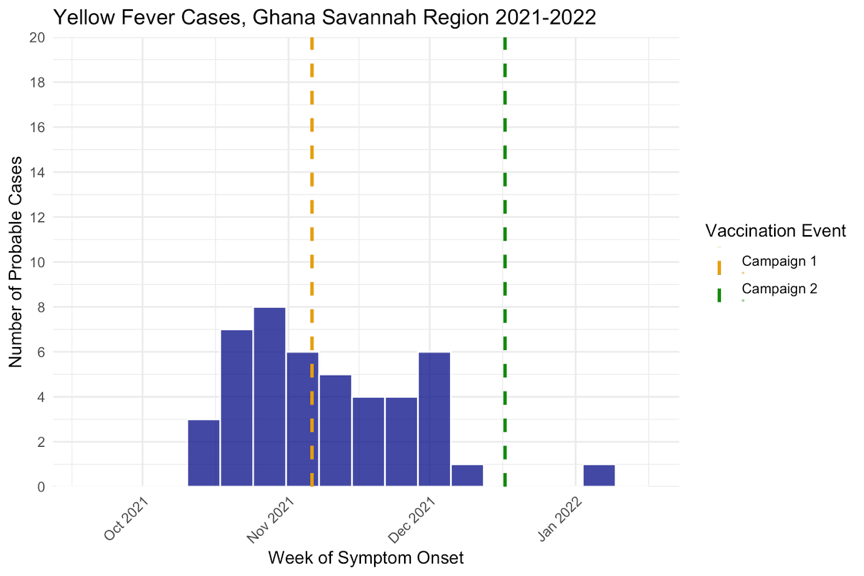

Figure S2. Ghana 2021-2022 outbreak epidemiological curve

The epidemiological curve of weekly cases from the 2021-2022 outbreak in the Savannah region of Ghana is shown along with the start dates of reactive vaccination campaigns.

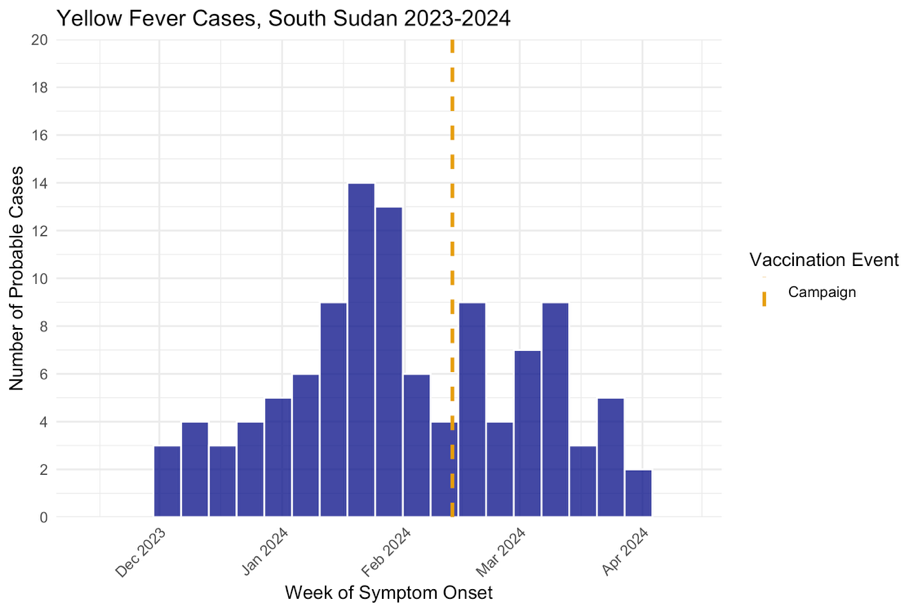

Figure S3. South Sudan 2023-2024 outbreak epidemiological curve

The epidemiological curve of weekly cases from the 2023-2024 outbreak in the South Sudan Western Equatoria regions is shown along with the start dates of the reactive vaccination campaign.

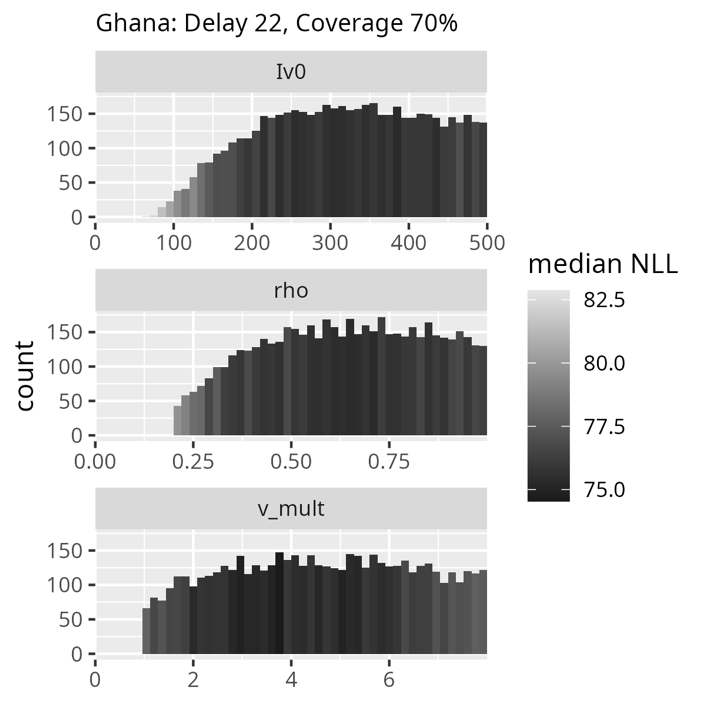

Figure S4. Distribution of estimated parameters for Ghana outbreak model calibration

Histograms represent the parameter sets for the estimated parameters of reporting ratio (rho*,*) initial infected mosquitoes (Iv), and mosquito-to-human ratio (v_mult) based on calibration to the Ghana 2021-2022 outbreak using negative log-likelihood (NLL). The panels show the Latin Hypercube sampling of all parameter sets within 10 NLL.

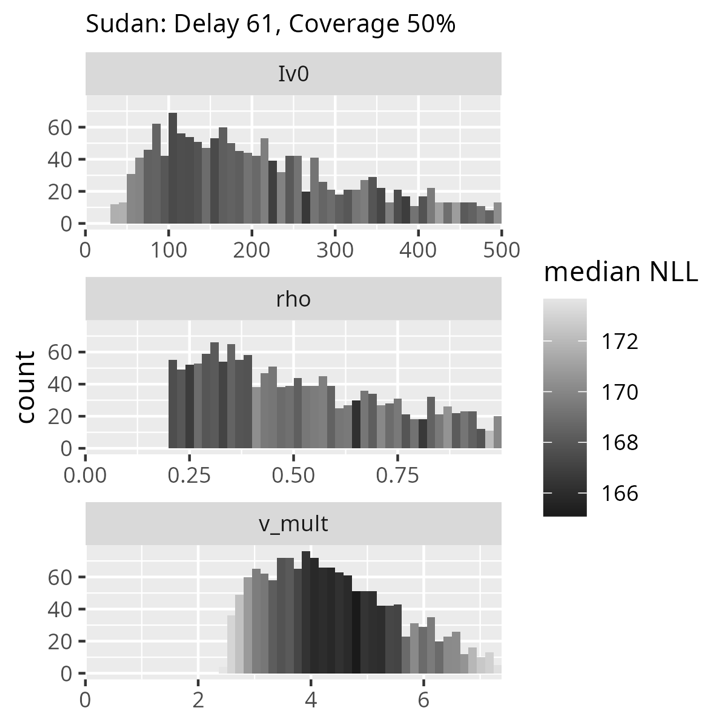

Figure S5. Distribution of estimated parameters for South Sudan outbreak model calibration

Histograms represent the parameter sets for the estimated parameters of reporting ratio (rho*,*) initial infected mosquitoes (Iv), and mosquito-to-human ratio (v_mult) based on calibration to the South Sudan 2023-2024 outbreak using negative log-likelihood (NLL). The panels show the Latin Hypercube sampling of all parameter sets within 10 NLL.

To create the model scenarios below, a likelihood-weighted sample was conducted on each of the parameter sets shown above. To determine the optimal value for the parameter sampling, we compared second-stage Monte Carlo convergence by repeated likelihood-weighted resampling of the retained first-stage parameter sets. At 4,400 draws the Monte Carlo standard error of the cumulative severe/symptomatic cases was 0.22% of the full weighted mean, corresponding to an approximate 95% resampling precision of ±0.43%. Therefore 4,400 was chosen as the sample size for the likelihood-weighted sampling.

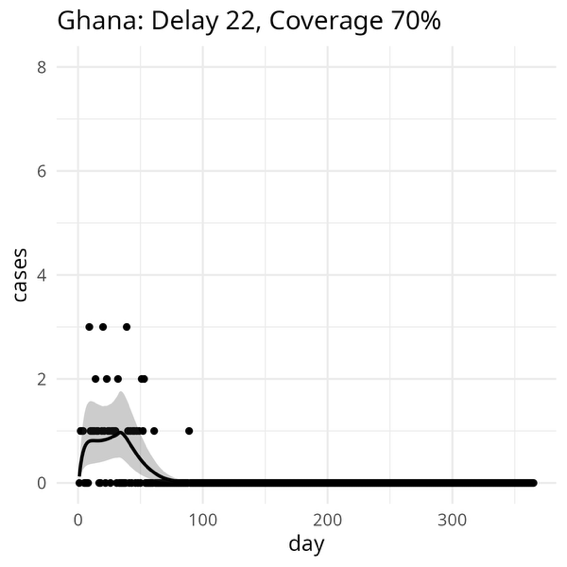

Figure S6. Observed vs modeled daily yellow fever cases in Ghana 2021-2022

The points depict the observed daily yellow fever cases in the Savannah region of Ghana from 2021-2022. The smooth black line and grey shaded region show the mean best fit and the 95% CI.

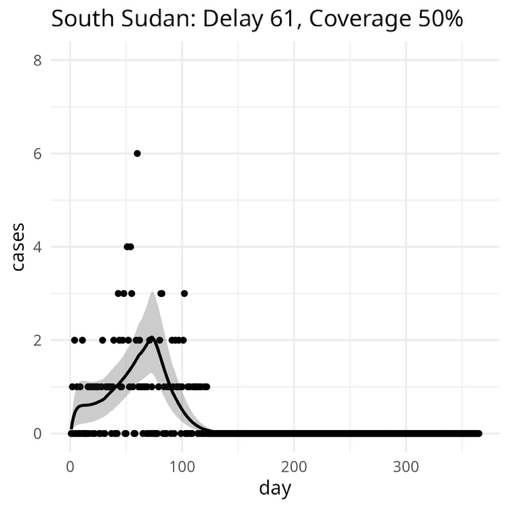

Figure S7. Observed vs modeled daily yellow fever cases in South Sudan 2023-2024

The points depict the observed daily yellow fever cases in South Sudan from 2023-2024. The smooth black line and grey shaded region show the mean best fit and the 95% CI.

4. Scenario Analysis

| Table S2. Ghana outbreak cases and deaths, observed vs simulated timing delays | | |
| --- | --- | --- |
| Scenario | Total cases | Total deaths |
| Observed (22-day delay) | 45 | 6 |
| Baseline scenario | 45 (32-58) | 7 (4-12) |
| 20-day delay | 42 (31-55) | 7 (4-11) |
| 60-day delay | 107 (62-177) | 17 (8-39) |
| 120-day delay | 326 (91-985) | 55 (12-203) |

| Table S3. Ghana outbreak cases and deaths, observed vs simulated vaccination coverage | | |
| --- | --- | --- |
| Scenario | Total cases | Total deaths |
| Observed (70% coverage) | 45 | 6 |
| Baseline scenario | 45 (32-58) | 7 (4-12) |
| 50% | 106 (73-149) | 17 (9-32) |
| 60% | 71 (51-96) | 11 (6-20) |
| 80% | 25 (18-33) | 4 (2-6) |

| Table S4. Ghana outbreak cases and deaths, observed vs simulated vaccination coverage and delays | | |
| --- | --- | --- |
| Scenario | Total cases | Total deaths |
| Observed (70%, 22-day delay) | 45 | 6 |
| Baseline scenario | 45 (32-58) | 7 (4-12) |
| 50%, 20-day delay | 98 (68-153) | 15 (8-29) |
| 50%, 60-day delay | 414 (159-910) | 68 (20-195) |
| 50%, 120-day delay | 2834 (303-9823) | 472 (39-1896) |
| 60%, 20-day delay | 66 (48-87) | 10 (6-18) |
| 60%, 60-day delay | 218 (104-424) | 35 (13-92) |
| 60%, 120-day delay | 1067 (172-3773) | 180 (22-758) |
| 80%, 20-day delay | 24 (17-31) | 4 (2-6) |
| 80%, 60-day delay | 46 (32-64) | 7 (4-14) |
| 80%, 120-day delay | 86 (43-178) | 14 (5-38) |

| Table S5. South Sudan outbreak cases and deaths, observed vs simulated timing delays | | |
| --- | --- | --- |
| Scenario | Total cases | Total deaths |
| Observed (61-day delay) | 110 | 6 |
| Baseline scenario | 111 (91-132) | 26 (11-52) |
| 20-day delay | 31 (23-41) | 7 (3-15) |
| 60-day delay | 108 (89-128) | 25 (11-51) |
| 120-day delay | 614 (380-940) | 141 (50-299) |

| Table S6. South Sudan outbreak cases and deaths, observed vs simulated vaccination coverage | | |
| --- | --- | --- |
| Scenario | Total cases | Total deaths |
| Observed (50%) | 110 | 6 |
| Baseline scenario | 111 (91-132) | 26 (11-52) |
| 50% | 111 (91-132) | 26 (11-52) |
| 60% | 63 (52-76) | 15 (6-30) |
| 80% | 16 (11-20) | 4 (2-7) |

| Table S7. South Sudan outbreak cases and deaths, observed vs simulated vaccination coverage and delays | | |
| --- | --- | --- |
| Scenario | Total cases | Total deaths |
| Observed (50%, 61-day delay) | 110 | 6 |
| Baseline scenario | 111 (91-132) | 26 (11-52) |
| 50%, 20-day delay | 31 (23-41) | 7 (3-15) |
| 50%, 60-day delay | 108 (89-128) | 25 (11-51) |
| 50%, 120-day delay | 614 (380-940) | 141 (50-299) |
| 60%, 20-day delay | 22 (16-29) | 5 (2-10) |
| 60%, 60-day delay | 62 (50-74) | 14 (6-29) |
| 60%, 120-day delay | 238 (166-331) | 55 (22-115) |
| 80%, 20-day delay | 8 (6-12) | 2 (1-4) |
| 80%, 60-day delay | 15 (11-20) | 4 (2-7) |
| 80%, 120-day delay | 27 (22-32) | 6 (3-13) |

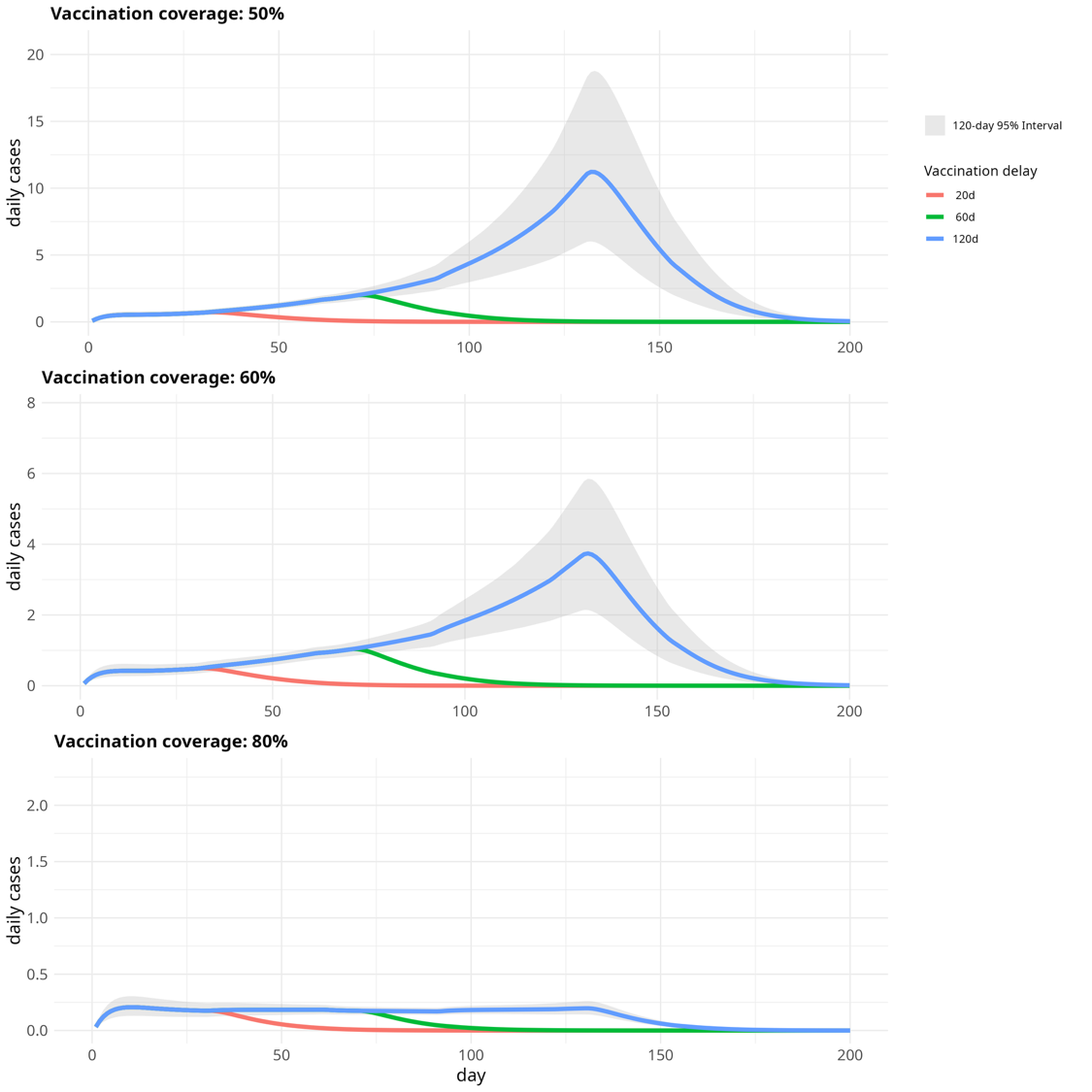

Figure S8. Projected yellow fever outbreak trajectories based on vaccination coverage and timing, South Sudan

The figure shows estimated severe/symptomatic yellow fever cases in counterfactual scenarios assuming vaccination coverage of 50%, 60%, and 80%. The red line in each panel indicates the estimated epidemic assuming a delay of 20 days to reactive vaccination, versus 60 days (green line) and 120 days (blue line). The shaded grey region represents the 95% uncertainty interval under the 120-day delay scenario (which overlaps with the other scenarios of vaccination delay until the time of simulated reactive vaccination).

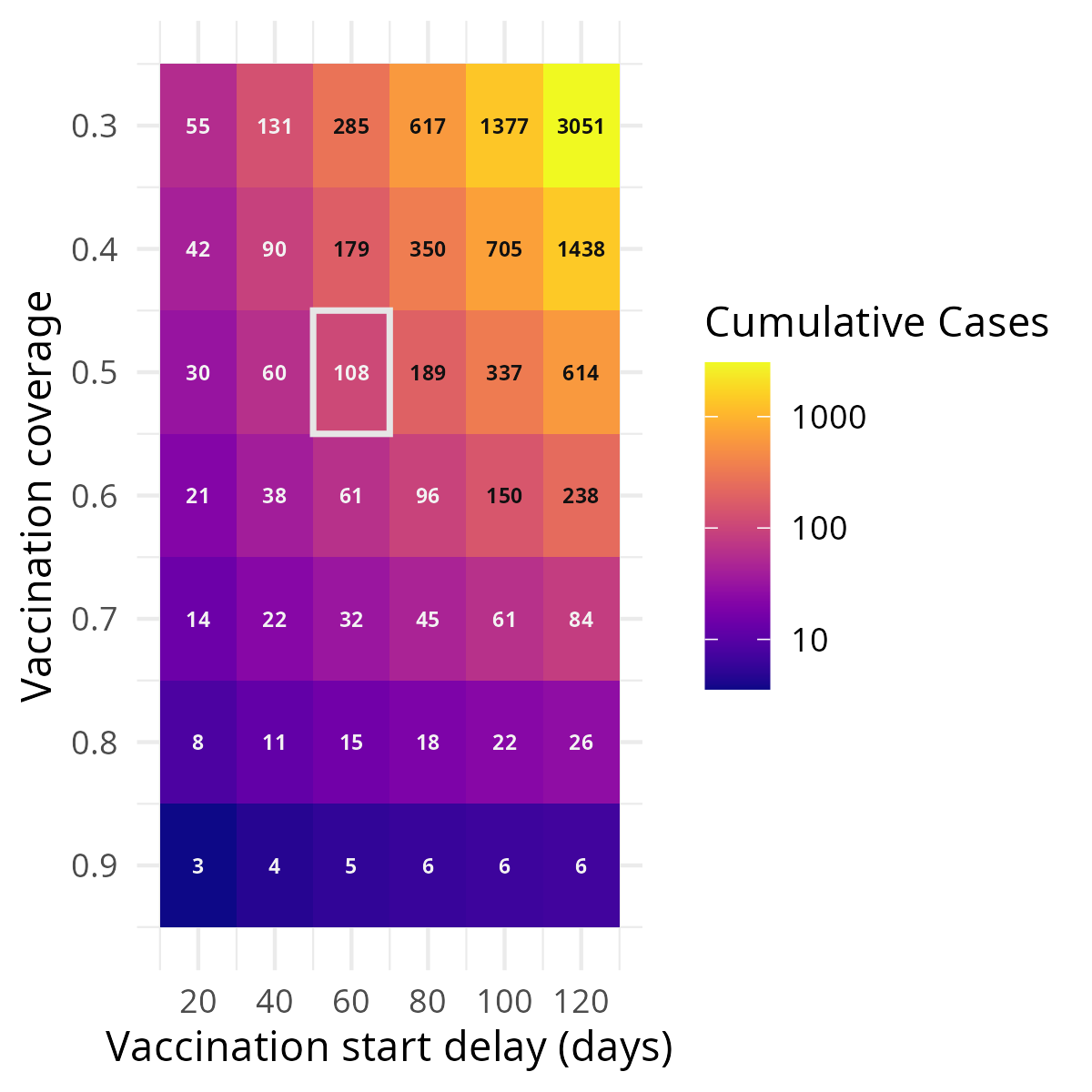

Figure S9. Heatmap of vaccination coverage vs timing on yellow fever outbreak size, South Sudan

The heatmap depicts the estimated number of cumulative symptomatic/severe yellow fever cases during the South Sudan 2023-2024 outbreak, under counterfactual scenarios of baseline vaccination coverage and delay to start of reactive vaccination. Color intensity indicates the mean number of estimated cases on a logarithmic scale. The scenario closest to the observed epidemic (50% baseline vaccination coverage, 61-day delay, 110 observed cases) is outlined in white*.*
